# RISK6, a universal 6-gene transcriptomic signature of TB disease risk, diagnosis and treatment response

**DOI:** 10.1101/19006197

**Authors:** Adam Penn-Nicholson, Stanley Kimbung Mbandi, Ethan Thompson, Simon C. Mendelsohn, Sara Suliman, Novel N. Chegou, Stephanus T. Malherbe, Fatoumatta Darboe, Mzwandile Erasmus, Willem A. Hanekom, Nicole Bilek, Michelle Fisher, Stefan H. E. Kaufmann, Jill Winter, Melissa Murphy, Robin Wood, Carl Morrow, Ildiko Van Rhijn, Branch Moody, Megan Murray, Bruno B. Andrade, Timothy R. Sterling, Jayne Sutherland, Kogieleum Naidoo, Nesri Padayatchi, Gerhard Walzl, Mark Hatherill, Daniel Zak, Thomas J. Scriba, the Adolescent Cohort Study team, GC6-74 Consortium, The ScreenTB and AE-TBC teams, CAPRISA IMPRESS team, RePORT Brazil Consortium, Peruvian Household Contacts Cohort study group

**Affiliations:** South African Tuberculosis Vaccine Initiative, Institute of Infectious Disease and Molecular Medicine and Division of Immunology, Department of Pathology, University of Cape Town, Cape Town, South Africa; Center for Infectious Disease Research, Seattle, WA, USA; Brigham and Women’s Hospital, Division of Rheumatology, Immunity and Inflammation, Harvard Medical School, Boston, USA; DST-NRF Centre of Excellence for Biomedical Tuberculosis Research; South African Medical Research Council Centre for Tuberculosis Research; Division of Molecular Biology and Human Genetics, Faculty of Medicine and Health Sciences, Stellenbosch University, Cape Town, South Africa; Max Planck Institute for Infection Biology, Berlin, Germany; Hagler Institute for Advanced Study at Texas A&M University, College Station, TX, USA; Catalysis Foundation for Health, San Ramon, CA, USA; Desmond Tutu HIV Centre, and Institute of Infectious Disease and Molecular Medicine (IDM), University of Cape Town, Cape Town, South Africa; Department of Global Health and Social Medicine, and Division of Global Health Equity, Brigham and Women’s Hospital, Harvard Medical School, Boston, MA, USA; Instituto Gonçalo Moniz, Fundação Oswaldo Cruz, Salvador, Brazil; Division of Infectious Diseases, Department of Medicine, Vanderbilt University School of Medicine, Nashville, USA; Vaccines and Immunity, Medical Research Council Unit, Fajara, the Gambia; Centre for the AIDS Programme of Research in Africa, Durban, South Africa; South African Medical Research Council-CAPRISA HIV-TB Pathogenesis and Treatment Research Unit, Durban, South Africa

## Abstract

Improved tuberculosis diagnostics and tools for monitoring treatment response are urgently needed. We developed a robust and simple, PCR-based host-blood transcriptomic signature, RISK6, for multiple applications: identifying individuals at risk of incident disease, as a screening test for subclinical or clinical tuberculosis, and for monitoring tuberculosis treatment. RISK6 utility was validated by blind prediction using quantitative real-time (qRT) PCR in seven independent cohorts.

Prognostic performance significantly exceeded that of previous signatures discovered in the same cohort. Performance for diagnosing subclinical and clinical disease in HIV-uninfected and HIV-infected persons, assessed by area under the receiver-operating characteristic curve, exceeded 85%. As a screening test for tuberculosis, the sensitivity at 90% specificity met or approached the benchmarks set out in World Health Organization target product profiles for non-sputum-based tests. RISK6 scores correlated with lung immunopathology activity, measured by positron emission tomography, and tracked treatment response, demonstrating utility as treatment response biomarker, while predicting treatment failure prior to treatment initiation. Performance of the test in capillary blood samples collected by finger-prick was noninferior to venous blood collected in PAXgene tubes. These results support incorporation of RISK6 into rapid, capillary blood-based point-of-care PCR devices for prospective assessment in field studies.

## INTRODUCTION

The “End Tuberculosis Strategy” of the World Health Organization (WHO) aims to reduce the annual incidence of tuberculosis (TB) to less than 10 cases per 100,000 people by the year 2035^1^. To achieve this goal the primary proposed strategy is to increase and improve efforts to find and treat individuals with active TB disease, to conduct universal screening of those at high risk, and to provide preventive therapy to those at risk of progressing to active TB disease^1^. There is thus a need for improved prognostic and diagnostic tests to identify those at risk of incident TB and those with subclinical or active TB, for appropriate treatment. The provision and management of TB treatment, as well as monitoring a patient’s response to treatment, also require much improvement. The standard 6-month regimen of treatment appears to be unnecessarily long for many patients with drug-susceptible TB, while insufficient to cure some patients, even in clinical trials when treatment adherence is maximized^2^. Experimental regimens tested in recent clinical trials have also been inadequate to cure treatment-refractory patients^3^. Collectively, these data support the now accepted principle that TB exists in a pathophysiological spectrum that spans several stages of infection, subclinical and active disease, including distinct stages of treatment outcome. Achieving the “End Tuberculosis Strategy” clearly depends on approaches that can place an individual into the stage of this spectrum such that clinical management is appropriate. A universal, non-sputum biomarker capable of predicting progression to active TB, diagnosing disease and monitoring the response to TB treatment would be a major advance in the efforts to achieve the “End Tuberculosis Strategy”. We hypothesized that a single, parsimonious host-blood transcriptomic signature can be developed for all three purposes with performance criteria that meet the target product profiles for tests to predict TB progression^4^ and for a TB screening test^5^ proposed by the WHO.

We sought to discover and validate a parsimonious and robust blood transcriptomic signature with universal applicability for predicting incident TB, as a triage test for identifying those who should be further investigated for TB disease, and for monitoring of TB treatment response. We explicitly set out to develop this signature for ultimate translation to a hand-held point-of-care platform and therefore performed all analyses, including signature training and performance assessments in all validation cohorts, by quantitative RT-PCR, using a highly standardized protocol and locked-down analysis algorithm. We assessed performance of RISK6 by blind prediction as a prognostic test for incident TB, as a TB diagnostic in HIV-uninfected and HIV-infected individuals, including individuals presenting with symptoms requiring investigation for TB at primary health care centres, and as a treatment response biomarker. We also tested the robustness of RISK6 and report performance of RISK6 measured in capillary blood samples collected by finger-prick, facilitating the way for incorporation into point-of-care diagnostic devices.

## RESULTS

### Prognostic performance of RISK6 in the Adolescent Cohort Study discovery cohort

The RISK6 signature was discovered on samples from adolescent progressors and controls (**Supplementary Figure 1a**) by selecting the smallest set of transcripts with the best prognostic performance based on qRT-PCR data, and comprises an ensemble of 9 transcript pairs formed between three transcripts upregulated in progressors (GBP2, FCGR1B, and SERPING1), and three transcripts downregulated in progressors (TUBGCP6, TRMT2A and SDR39U1), relative to non-progressors (**Supplementary Table 1 and Figure 1a-b**). We first sought to determine if the prognostic performance of RISK6 for incident TB in the discovery cohort was comparable to that of the previously published ACS 16-gene signature^6^, consisting of 57 transcripts (PCR primer/probe assays) or the ACS 11-gene version (48 PCR primer/probe assays), which was developed for greater throughput in multiplex assays^7^. The PCR-based RISK6 and both ACS signatures readily discriminated between Adolescent Cohort Study progressor and non-progressor samples collected within 12 months of TB diagnosis (**Figure 1c** and **Supplementary Table 2**). Interestingly, prognostic performance of RISK6, estimated by model fit (AUC 87.6%, 95%CI 82.8 - 92.4), was significantly better than ACS 16-gene (AUC 81.8%, 95%CI 75.1 - 88.6, pROC analysis p = 0.024) and ACS 11-gene (AUC 82.2%, 95%CI 75.6 - 88.8, pROC analysis p = 0.03). As observed previously with the 16-gene signature^6^, RISK6 also discriminated between progressors and non-progressors using samples collected between 12 and 24 months before TB disease diagnosis (AUC 74.0%, 95%CI 66.0 - 82.0), although discrimination was weaker than observed on samples within a year of diagnosis (**Figure 1d)**.

**Table 1:**
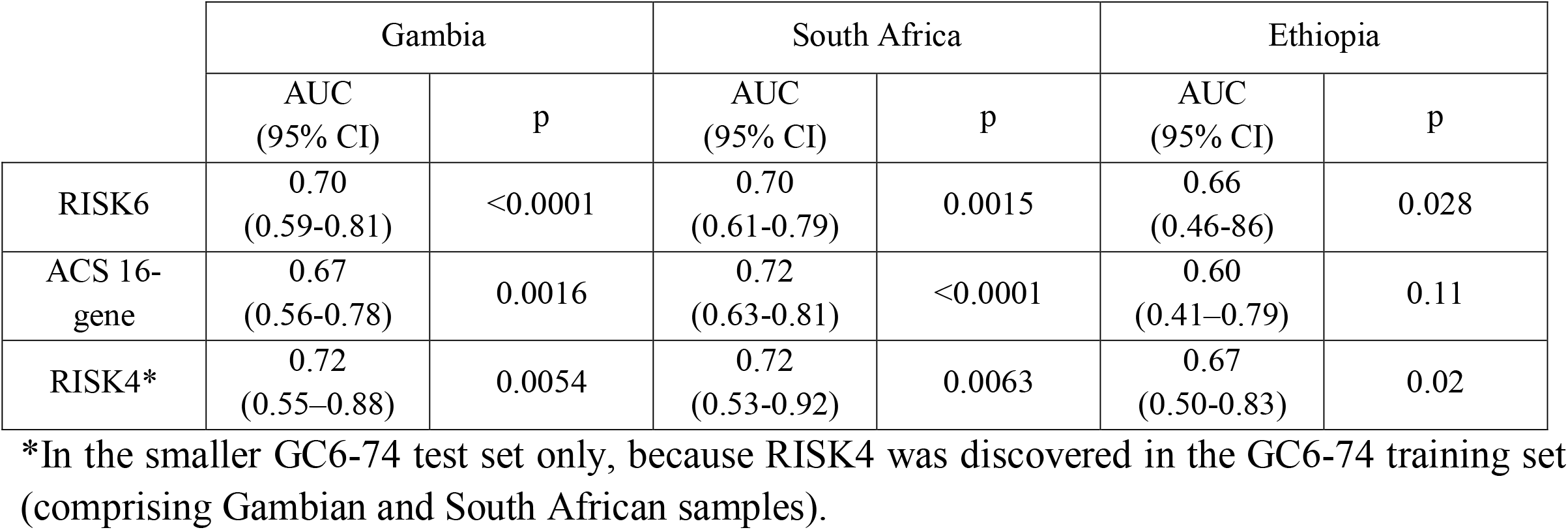
Performance of RISK6 signature in the GC6 cohort, compared to the ACS 16-gene and RISK4 signatures, by blinded validation. Samples from 0-24 months before TB diagnosis were included.

**Table 2:**
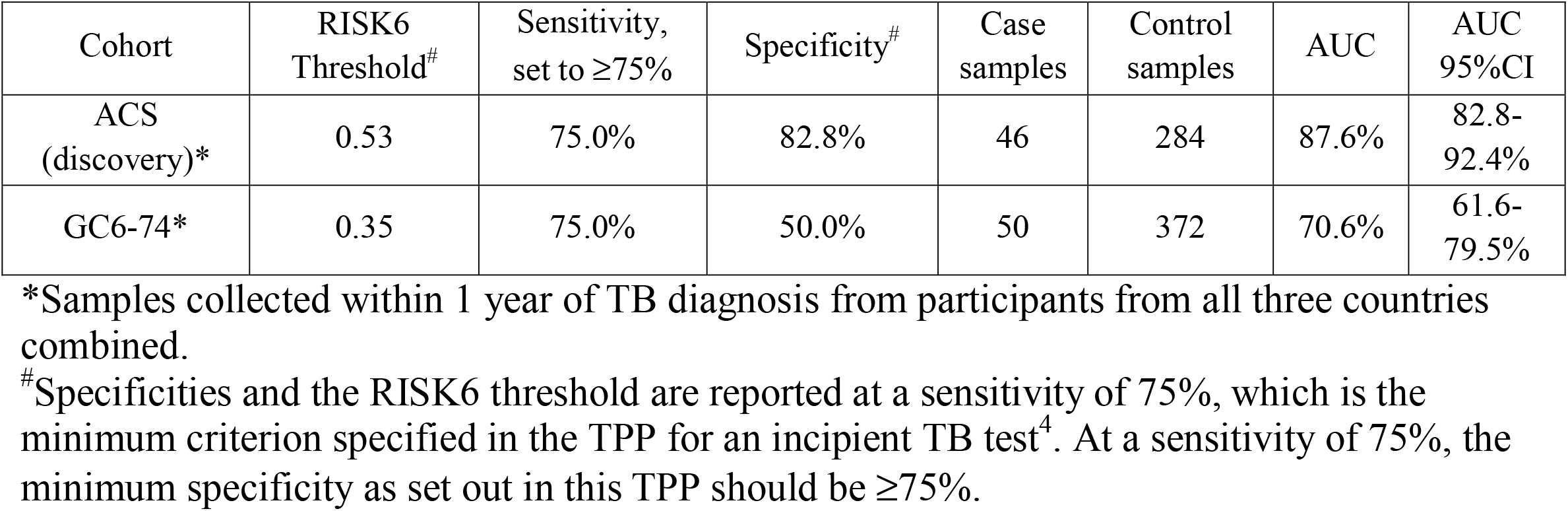
Accuracy of the RISK6 signature benchmarked against the WHO target product profile for prediction of incident TB

**Figure 1:**
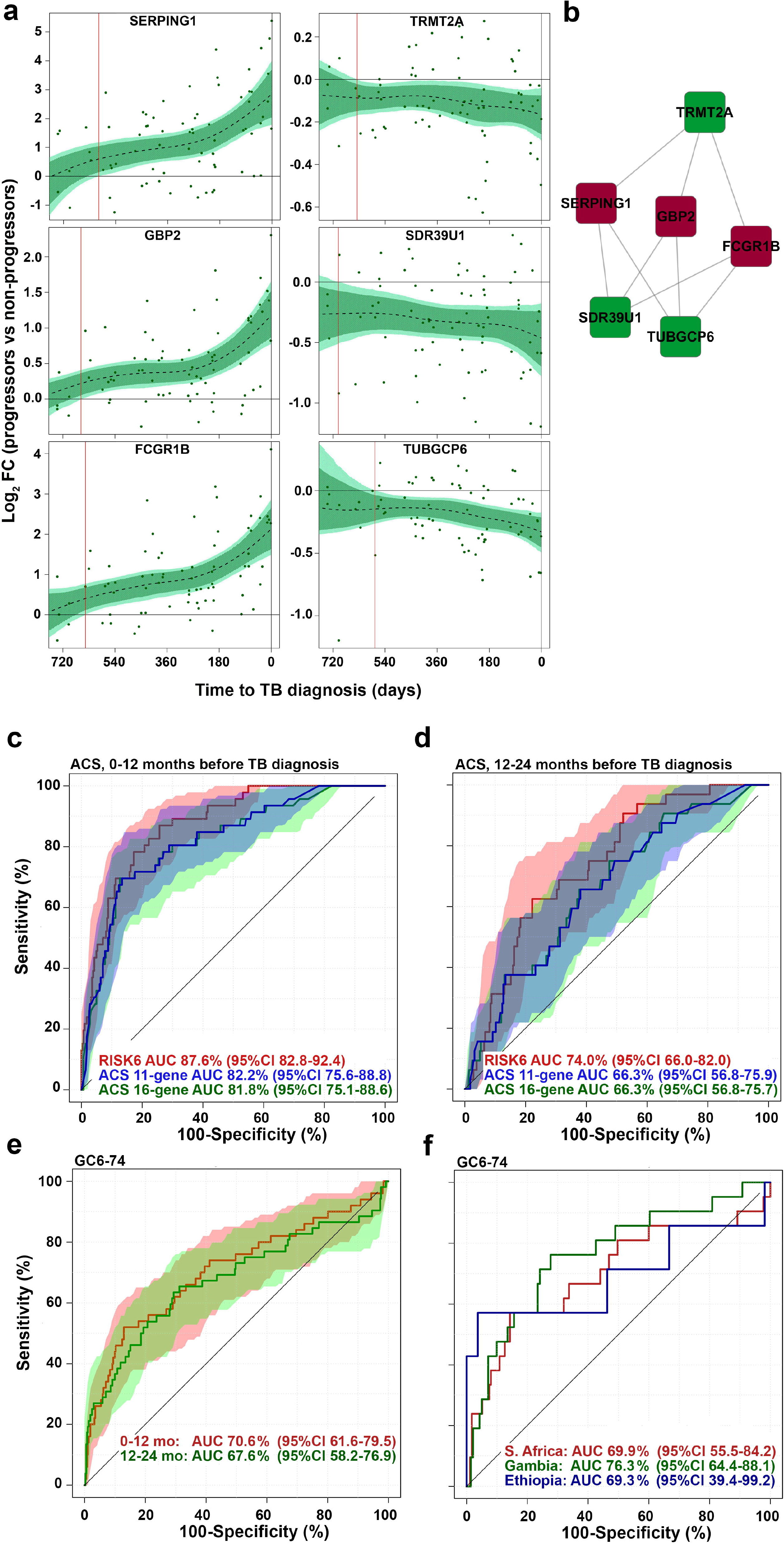
Discovery of the RISK6 signature. (a) Expression kinetics of the six transcripts in RISK6 signature over time, measured by RNA-sequencing and expressed as log_2_ fold change between matched adolescent progressors and controls and modelled as non-linear splines (dotted lines). Light blue shading 95% CI for the temporal trends, computed by performing 2000 spline fitting iterations after bootstrap resampling from the full dataset. Transcripts that are upregulated during TB progression are on the left and those that are downregulated and on the right. (b) RISK6 comprises nine pairs that each link a transcript that is upregulated during TB progression with one that is downregulated, relative to healthy controls. Lines indicate pairing (refer to Table 1 for TaqMan primer-probe sets that match the transcripts). Transcripts that are upregulated in progressors are in red nodes and those that are downregulated in progressors are in green. (c and d) Receiver operating characteristic (ROC) curves depicting the performance (model fit) of RISK6, relative to the ACS 11-gene^7^ and ACS 16-gene^6^ signatures, measured by qRT-PCR on RNA from whole blood samples collected from participants of the ACS cohort within one year of TB disease diagnosis (c), or 1-2 years before TB diagnosis (d). Shaded areas depict the 95% CI. (e) Receiver operating characteristic (ROC) curves depicting prognostic performance, by blind prediction of RISK6, measured by qRT-PCR on RNA from whole blood RNA collected from participants of the GC6-74 cohort of household contacts. Shaded areas depict the 95% CI. (f) Prognostic performance of RISK6 for incident TB in household contacts from South Africa, The Gambia or Ethiopia, measured by qRT-PCR on RNA from whole blood RNA collected within 1 year of TB diagnosis from participants of the GC6-74 cohort. Performance on 0-2 years before TB diagnosis is shown in Table 1.

### Validation of RISK6 prognostic performance in the GC6-74 cohort

We validated prognostic performance of the RISK6 signature for incident TB by blind prediction on the independent GC6-74 cohorts of household TB contacts from South Africa, The Gambia and Ethiopia, who either progressed to TB or remained asymptomatic^8^. RISK6 significantly discriminated between GC6-74 progressors and non-progressors on samples collected within 12 months of incident TB diagnosis (AUC 70.6%, 95%CI 61.6 - 79.5) and those collected 12-24 months before TB diagnosis (AUC 67.6%, 95%CI 58.2 - 76.9, **Figure 1e** and **Table 1**). At a sensitivity threshold of 75%, RISK6 achieved a specificity of 50.3% within 1 year of diagnosis in the GC6-74 cohort, which does not meet the WHO target product profile for a test predicting progression from tuberculosis infection to active disease^4^ (**Table 2** and **Supplementary Table 2**).

Since RISK6 was discovered on a South African cohort it was important to determine if performance varies by geography, since differences in population genetic structure, local epidemiology and environment may influence blood biomarker performance^9^. We therefore also assessed prognostic performance by country. Interestingly, when assessing samples collected within 12 months of TB diagnosis, the AUC was highest for the Gambian cohort (AUC 76.3%, 95%CI 64.4% - 88.1%, **Figure 1f** and **Table 1**), while the AUC for the South African cohort was similar to the entire, combined GC6-74 cohort (AUC 69.9%, 95%CI 55.5 - 84.2, **Figure 1f**). Although the AUC for the smaller Ethiopian cohort (comprising 12 progressors) was also similar (AUC 69.3%, 95%CI 39.4 - 99.2), the confidence intervals were very large and discrimination between progressors and non-progressors was not significant (**Figure 1f**). RISK6 also validated on samples from the entire 24 month period prior to TB diagnosis from these 3 GC6-74 cohorts (**Table 1**).

### Performance of RISK6 as a screening test in HIV-uninfected and HIV-infected individuals

Expression levels of the six transcripts in RISK6 differed most between adolescent progressors and non-progressors at the time of TB diagnosis (**Figure 1a**). In light of this, we hypothesized that RISK6 would also yield good performance as a screening or triage test for TB. Since HIV infection is a major risk factor for TB and a large proportion of TB patients in settings endemic for TB are HIV-infected^10^, we aimed to determine diagnostic performance in both HIV-infected and uninfected individuals. We therefore compared diagnostic performance of RISK6, benchmarked against the ACS 11-gene signature, in 112 HIV-uninfected (61 asymptomatic controls and 51 TB cases) and 82 HIV-infected (40 asymptomatic controls and 42 TB cases) adults from the Western Cape, South Africa. Excellent diagnostic performance of RISK6 was seen in both HIV-uninfected (AUC 93.7%, 95%CI 87.9-99.4%) and HIV-infected persons (AUC 92.6%, 95%CI 86.8-98.5); performance was not different between the two groups (pROC analysis p = 0.76, **Figure 2a**). By contrast, diagnostic performance of the ACS 11-gene signature was better in HIV-uninfected (AUC 97.3%, 95%CI 93.7-100) than in HIV-infected persons (AUC 87.9%, 95% CI 80.6-95.2); the 9% lower AUC in HIV-infected persons was significant (pROC analysis p = 0.027, **Figure 2b**)RISK6 signature scores were higher in HIV-infected controls compared to HIV-uninfected controls, suggesting an effect of underlying HIV infection on RISK6 (**Figure 2c**). To understand the effects of underlying HIV infection on the RISK6 signature, we determined the difference in expression of each transcript between HIV-infected and uninfected individuals. Expression of FCGR1B and GBP2, but none of the other transcripts, was significantly higher in HIV-infected than uninfected controls, while no significant differences were observed in TB cases (**Figure 2d**). At a sensitivity threshold of 90%, RISK6 achieved a specificity of 93.4% and 72.5% in HIV-uninfected and HIV-infected persons, respectively (**Table 3** and **Supplementary Table 2**).

**Table 3:**
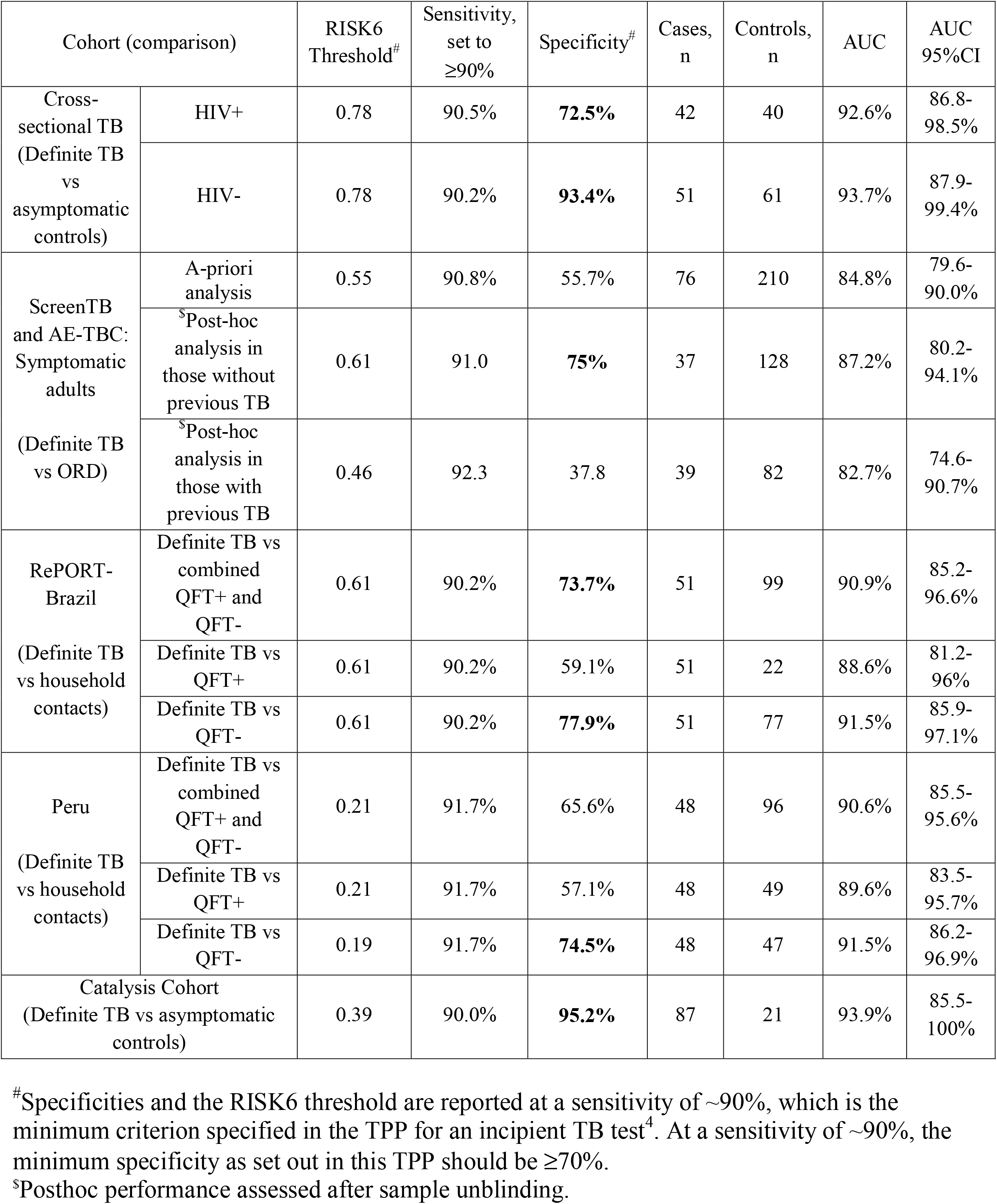
Accuracy of the RISK6 signature benchmarked against the WHO target product profile for a screening/triage test

**Figure 2:**
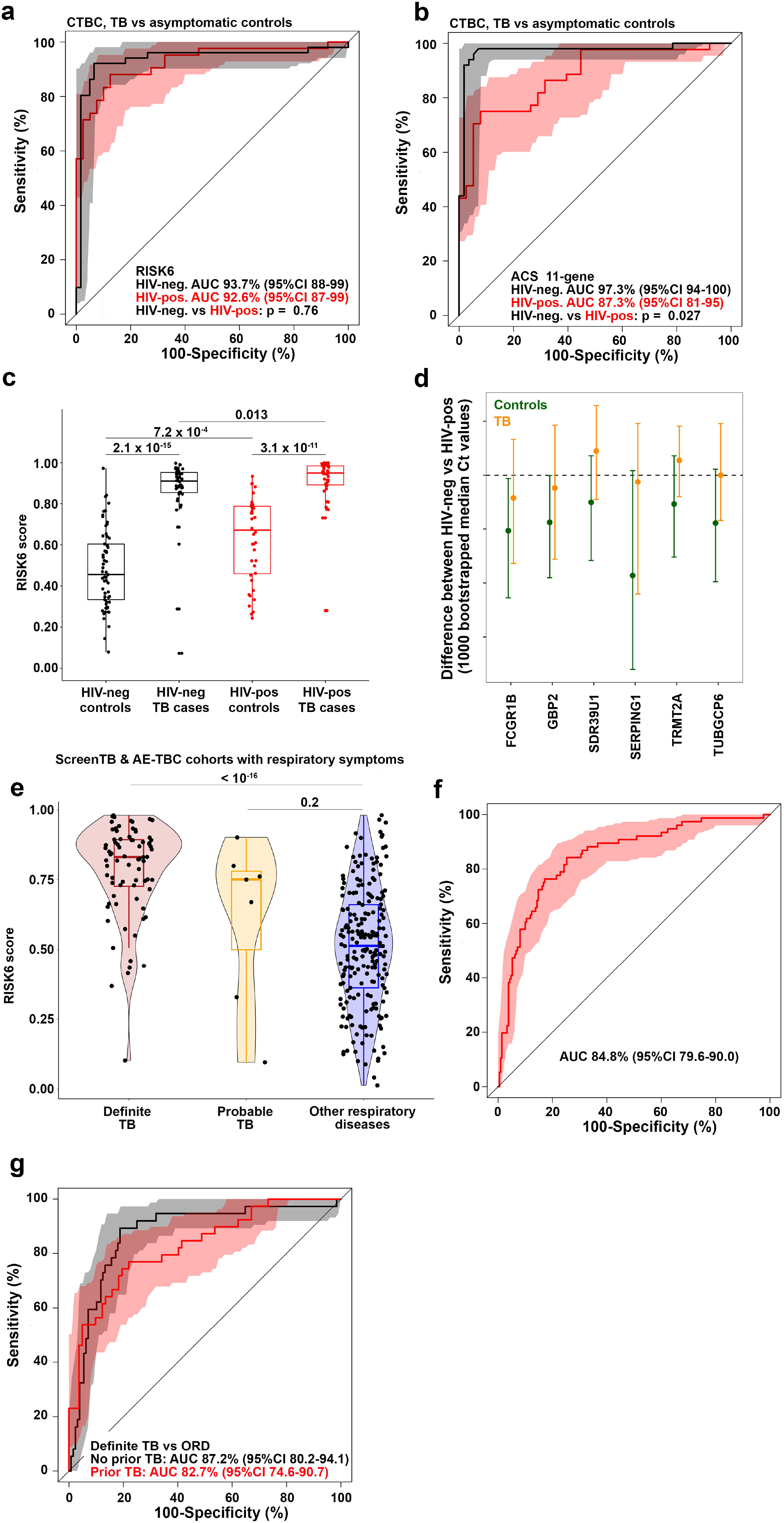
Diagnostic performance of RISK6 as a triage test. (a and b) ROC curves depicting diagnostic performance of RISK6 (a) and the ACS 11-gene signature (b), for discrimination between active TB cases and *Mtb*-infected controls in HIV-negative or HIV-positive individuals. RISK6 was measured by qRT-PCR on RNA from whole blood RNA. (c) Comparison of RISK6 signature scores in *Mtb*-infected controls and active TB cases in HIV-negative or HIV-positive individuals. P-values were calculated using the Mann-Whitney U test. Horizontal lines represent medians; boxes represent the IQR and whiskers the range. Dots represent individual sample scores. (d) Relative differences in RISK6 transcript expression levels between HIV-negative and HIV-positive *Mtb*-infected controls (green) or active TB cases (orange). Dots depict medians and error bars the 95% CI, calculated from 1000 bootstrapped Ct values. The dashed line represents zero. (e) Comparison of RISK6 signature scores, by blind prediction, in patients with definite TB (with rigorous microbiological conformation), patients with probable TB and in patients with other respiratory diseases (ORD). Horizontal lines depict medians, boxes the IQR and the whiskers the range. Violin plots depict the density of data points. P-values were computed by Mann-Whitney U test. (f) Receiver operating characteristic (ROC) curve depicting discrimination between prognostic performance, by blind prediction of RISK6, measured by qRT-PCR in definite TB patients and patients with other respiratory diseases (ORD). Shaded areas depict the 95% CI. (g) ROC curves depicting RISK6 discrimination between definite TB and ORD participants of the ScreenTB and AE-TBC cohorts stratified into participants with no history of prior TB (black), or in those with a history of prior TB (red). Shaded areas depict the 95% CI.

### Diagnostic performance of RISK6 as a screening test in patients with respiratory symptoms

We also determined RISK6 performance as a screening test in symptomatic adults enrolled into the ScreenTB ^11^ and AE-TBC studies^12,13^. These adult participants presented at primary health care clinics in Cape Town, South Africa with symptoms requiring investigation for TB including coughing for >2 weeks and at least another symptom consistent with TB, and were enrolled prior to the establishment of a TB or other disease diagnosis. RISK6 signature scores were measured by blind prediction on PAXgene blood collected at presentation for care before treatment initiation in (1) 76 patients with microbiologically-confirmed, definite TB, (2) 7 patients with probable TB, and (3) 210 patients with other respiratory diseases (ORD) (**Figure 2e**, see **Supplementary Table 4** for diagnostic criteria). RISK6 discriminated between definite TB and ORD patients with an AUC of 84.8% (95%CI 79.6-90.0, **Figure 2f**). At a sensitivity threshold of 90%, RISK6 achieved a specificity of 57.1% (95%CI 32.9-76.7) in these symptomatic patients (**Table 3** and **Supplementary Table 2**), which falls short of the WHO target product profile for a community-based triage or referral test to identify people suspected of having TB^5^. RISK6 performance did not differ between HIV positive and negative participants (HIV-neg, n = 250: AUC 85.4%, 95%CI 79.7-91.0; HIV-pos, n = 36: AUC 79.5%, 95%CI 65.1-94.0, data not shown).

We unpacked the performance of RISK6 further in posthoc analyses performed after unblinding of patient diagnostic status. We noted that a considerable number (n = 121) of ScreenTB and AE-TBC participants had a record of least one previous episode of TB; this is typical of patients presenting for TB investigation in such high-incidence settings ^10^. RISK6 discriminated between definite TB and ORD among participants with no history of prior TB with an AUC of 87.2% (95%CI 80.2-94.1%), whereas in those with a history of prior TB the AUC was 82.7% (95%CI 74.6-90.7%, **Table 3, Figure 2g**). Although these AUCs were not statistically different, the specificities at a set sensitivity of >90% were markedly different at 75.0% and 37.8%, respectively (**Table 3**).

We also applied RISK6 to published microarray datasets, using a risk score algorithm, RISK6geo, adapted for application to microarray or RNA-seq data. RISK6geo discriminated between TB cases and asymptomatic M.tb-infected controls with AUCs exceeding 90% in all cohorts (**Supplementary Table 3**). Comparative performance characteristics of RISK6 and RISK6geo on qRT-PCR data from the different cohorts in this study are also shown in **Supplementary Table 2**.

### Performance of RISK6 as a TB treatment monitoring biomarker

Diagnostic performance of RISK6 was also assessed in the Catalysis cohort of TB patients who were studied during and after TB treatment^14-16^. RISK6 achieved an AUC of 93.5 (95%CI 85.5-100) for discriminating between newly diagnosed TB cases and asymptomatic controls (**Figure 3a**). Next, we determined if RISK6 has utility as a biomarker for monitoring TB treatment. We hypothesized that RISK6 scores, which are very high in patients with active disease, would decrease rapidly during TB treatment such that samples collected after bacteriological cure can be discriminated from the respective pre-treatment sample with high accuracy. We also hypothesized that RISK6 would allow discrimination of cured patients from those with treatment failure after 24 weeks of treatment. When measured by qRT-PCR in patients with bacteriological cure in the Catalysis cohort, RISK6 scores decreased significantly during TB treatment, although scores observed at the end of treatment were still significantly higher than those observed in healthy controls (**Figure 3b**), as reported for the 16-gene ACS signature previously^16^. Despite this, RISK6 significantly discriminated between samples collected pre-treatment and one week after treatment initiation (AUC 79.5%, 95% CI 72.2-86.7), four weeks after treatment initiation (AUC 77.4%, 95% CI 69.9-84.9) and end of treatment samples (AUC 88.1%, 95% CI 82.5-93.6; (**Figure 3c**). Importantly, RISK6 was a strong predictor of treatment outcome and significantly differentiated between the 78 patients with bacteriological cure and the 7 patients with treatment failure at the time of TB diagnosis (AUC 77.1, 95% CI 52.9-100, **Figure 3d**), and at the end of treatment (AUC 95.2, 95% CI 87.5-100, **Figure 3d**). These data are consistent with RISK6 detecting differences in inflammatory profiles before the initiation of treatment which predict the outcome of treatment, while also detecting ongoing inflammation in those who fail treatment and do not achieve bacteriological cure by 24 weeks. To address this further, we determined if blood RNA signature scores were associated with *in vivo* pulmonary inflammation measured by ^18^F-labeled fluorodeoxyglucose (^18^F FDG) PET-CT. Surprisingly, RISK6 scores directly correlated metabolic activity in lung lesions as measured by total glycolytic activity index (TGAI) (Spearman ρ = 0.66, p < 0.0001, **Figure 3e**), while signature scores correlated inversely with Xpert Ct values (Spearman ρ = −0.60, p < 0.0001) and Mycobacterial Growth Indicator Tube (MGIT) culture days-to-positivity values (Spearman ρ = −0.67, p < 0.0001) (data not shown).

**Figure 3.**
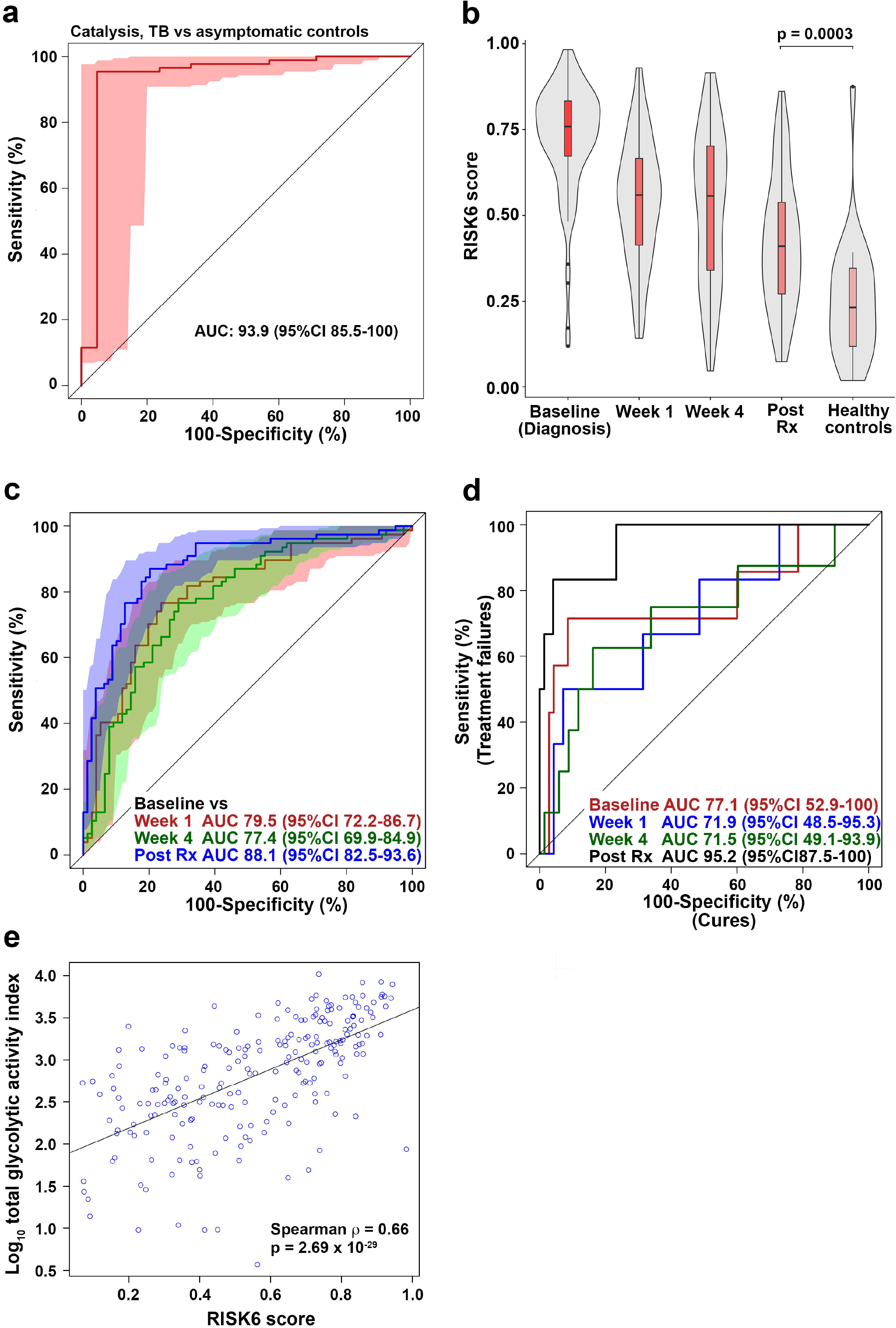
Treatment monitoring using the RISK6 signature in the Catalysis TB Treatment Cohort. (a) ROC curve depicting diagnostic performance of RISK6 for discriminating between active TB cases (irrespective of treatment outcome), sampled prior to treatment initiation (baseline, n=87), and controls (n=21) from the Catalysis Cohort. Shaded areas depict the 95% CI. (b) Comparison of RISK6 signature scores in cases (irrespective of treatment outcome) from the Catalysis Cohort at baseline and week 1 or week 4 after treatment initiation and after treatment completion (Post Rx). Also shown are the RISK6 signature scores in the healthy controls. Horizontal lines depict medians, the boxes the IQR and the whiskers the range. Violin plots depict the density of data points. The p-value, computed by Mann-Whitney U test, compares RISK6 signature scores after treatment completion with those in controls. (c) ROC curves depicting performance of RISK6 for discriminating between baseline (pre-treatment) samples and samples collected after week 1, week 4 or completion of TB treatment. (d) Prediction of treatment failure using RISK6. ROC curves depict discrimination between cases with cure (n = 70) and those with treatment failure (n = 7) in samples collected at treatment initiation (Baseline, red), at week 1 (blue), week 4 (green) and after treatment completion (Post Tx, black). (e) RISK6 scores plotted versus total glycolytic activity index measured by PET-CT for all available samples.

### Performance of RISK6 as triage test and TB treatment monitoring biomarker in South American Cohorts

An important issue is how a biosignature that was trained and validated in African cohorts will perform in geographically distinct populations. To address this, we assessed diagnostic performance of RISK6 measured by qRT-PCR in cohorts from Peru and Brazil. In the Peruvian cohort, RISK6 discriminated between culture-positive TB patients and QFT-negative with an AUC of 91.5% (95%CI 86.2-96.9, **Figure 4a**) and between TB patients and QFT-positive asymptomatic controls with an AUC of 89.6% (95%CI 83.5-95.7, **Figure 4b**). RISK6 also achieved an AUC of 90.9% (95%CI 85.2-96.6) for discriminating between Brazilian culture-positive TB patients and asymptomatic controls (**Figure 4c**). The minimum criteria for a screening or triage test for TB, set out by the WHO, set the sensitivity at ≥90% at a specificity of 70%^5^. With sensitivity set at ≥90%, the specificities for RISK6 applied to the South African HIV-negative (Cross-sectional and Catalysis cohorts) and HIV-positive cohorts far exceeded the criteria, with specificities above 90% (**Table 3** and **Supplementary Table 2**). Interestingly, performance of RISK6 in the South American cohorts also met these criteria when discriminating between TB cases and QFT-negative controls, but discrimination between TB cases and QFT-positive asymptomatic controls fell short of the 70% specificity mark (**Table 3** and **Supplementary Table 2**). It was notable that the RISK6 score threshold at which the sensitivity was ∼90% was quite variable, suggesting that the positivity cut-off for such a transcriptomic signature may be different for different cohorts.

**Figure 4.**
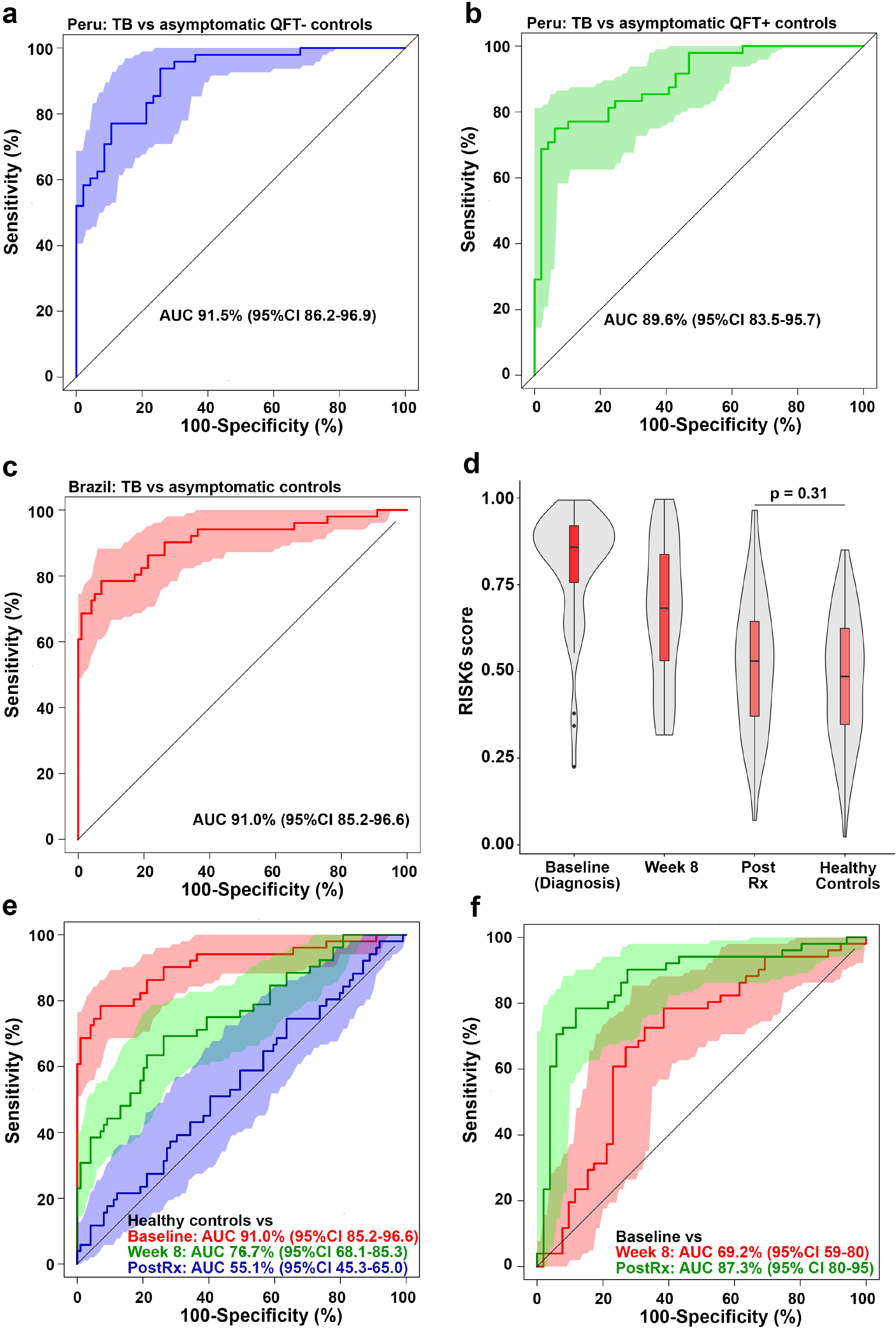
Diagnostic performance and treatment monitoring in South American cohorts. (a-b) ROC curve depicting diagnostic performance of RISK6 for discriminating between (a) culture-positive active TB cases (n = 48) and QuantiFERON-negative controls (n=47), or between (b) culture-positive active TB cases (n = 48) and QuantiFERON-positive controls (n=49) from the Peru Cohort. Shaded areas depict the 95% CI. (c) ROC curve depicting diagnostic performance of RISK6 for discriminating between culture-positive active TB cases, sampled prior to treatment initiation (baseline, n=51), and controls (n=99) from the RePORT-Brazil Cohort. (d) Comparison of RISK6 signature scores in TB cases at baseline, week 8 after treatment initiation and after treatment completion (Post Rx). Also shown are the RISK6 signature scores in healthy controls from Brazil. Horizontal lines depict medians, the boxes the IQR and the whiskers the range. Violin plots depict the density of data points. The p-value, computed by Mann-Whitney U test, compares RISK6 signature scores after treatment completion with those in controls. (e) ROC curves depicting performance of RISK6 for discriminating between healthy control samples and samples collected from TB cases before treatment initiation (baseline), at week 8 after treatment initiation, or completion of TB treatment (Post Rx). (f) ROC curves depicting performance of RISK6 for discriminating between baseline samples from TB cases and samples collected 8 weeks after treatment initiation, or upon completion of TB treatment (Post Rx).

We also determined performance of RISK6 as a biosignature for monitoring TB treatment in the Brazilian patients, all of whom achieved microbiological cure after 6 months of treatment.

RISK6 scores decreased significantly after 8 weeks of TB treatment and, unlike the Catalysis cohort, scores observed in the Brazilian patients at the end of treatment had reached levels observed in healthy controls (**Figure 4d and e**). RISK6 also significantly discriminated between samples collected pre-treatment and 8 weeks after treatment initiation (AUC 67.5%, 95% CI 56.4-78.6), and end of treatment samples (AUC 87.4%, 95% CI 79.8-94.9, **Figure 4f**).

### RISK6 as a treatment biomarker in HIV-infected patients with recurrent TB

The promising results from these treatment response studies prompted us to also evaluate if RISK6 can monitor success of recurrent TB treatment in HIV-infected individuals on ART, who participated in the randomized controlled IMPRESS trial. IMPRESS determined if a retreatment regimen that contained moxifloxacin, instead of ethambutol, would improve TB retreatment outcomes relative to the standard regimen^17^. No differences in RISK6 scores were observed between the two treatment arms of the trial (data not shown). Consequently, all analyses were performed with the treatment arms combined. RISK6 scores decreased upon treatment (**Figure 5a**) and could discriminate significantly between samples collected at the pre-treatment time point (baseline) and those collected after the intensive phase of treatment, at 2 months (AUC 75.1%, 95%CI 66.5-83.8, **Figure 5b**). Discrimination between baseline samples and those collected at the end of treatment, when all patients had achieved clinical cure, was better than after 2 months of treatment (AUC 91.2%, 95%CI 86.0-96.3, **Figure 5b**), although inflammation appeared to resolve further after the end of treatment, since RISK6 discriminated best between baseline and samples collected 6-8 months after treatment completion (AUC 98.5%, 95%CI 96.5-100, **Figure 5b**). When measured at baseline (not shown) or the end of the intensive treatment phase at 2 months (AUC 63.4%, 95%CI 48.2-78.5), RISK6 did not discriminate significantly between patients who had sputum culture conversion and those who converted after 2 months (**Figure 5c**). It was not possible to determine if RISK6 could predict treatment failure in this trial since all patients achieved bacteriological cure.

**Figure 5:**
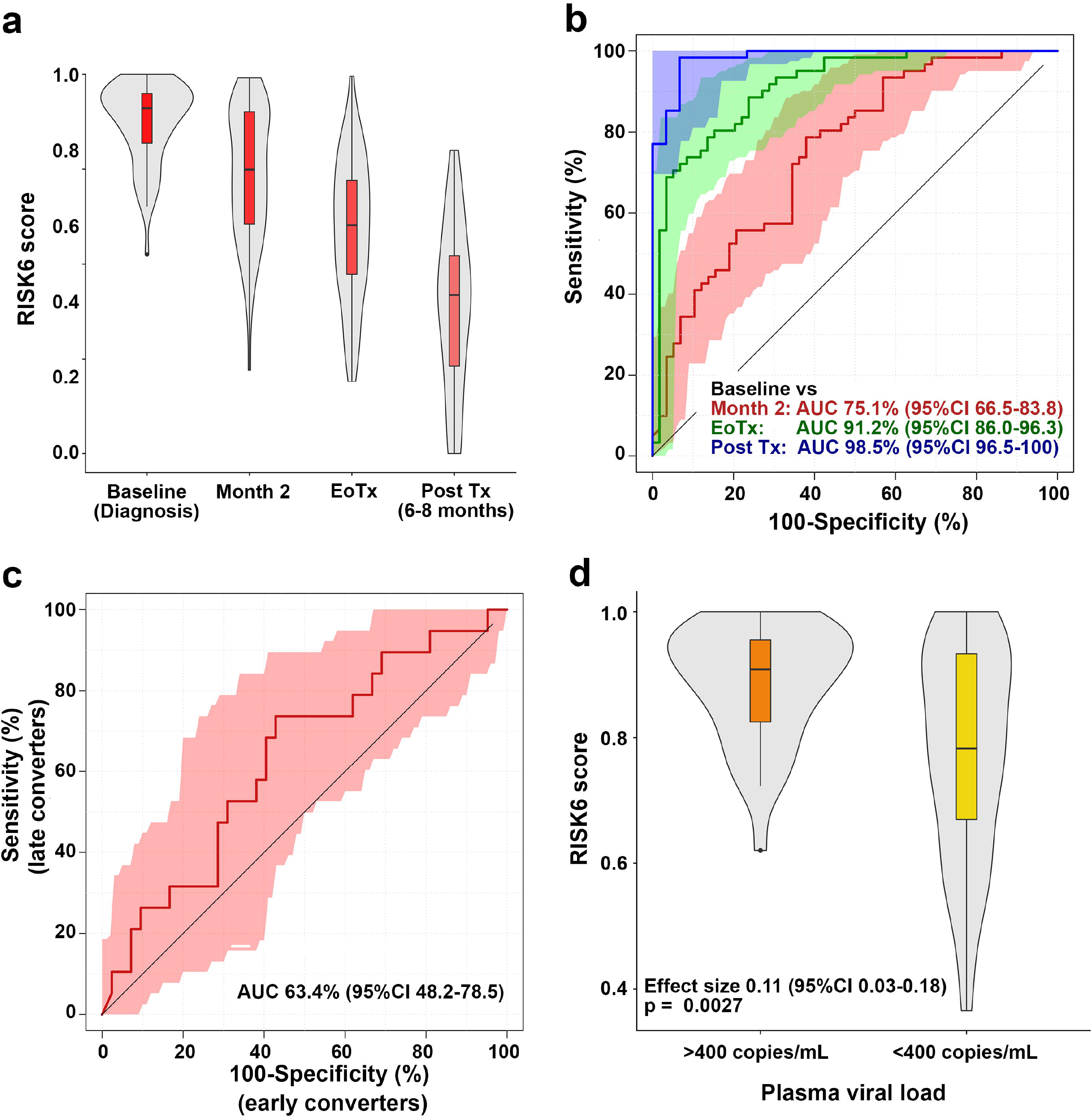
Treatment monitoring using the RISK6 signature in the IMPRESS cohort. (a) Comparison of RISK6 signature scores in HIV-infected TB cases (irrespective of treatment outcome) from the IMPRESS cohort at baseline, month 2 after treatment initiation, at treatment completion (EoRx) and 6-8 months after treatment completion (Post Rx). Horizontal lines depict medians, the boxes the IQR and the whiskers the range. Violin plots depict the density of data points. (b) ROC curves depicting performance of RISK6 for discriminating between baseline (pre-treatment) samples and samples collected at month 2 after treatment initiation, at treatment completion (EoRx) and 6-8 months after treatment completion (Post Rx). Shaded areas depict the 95% CI. (c) ROC curve depicting performance of RISK6, measured at 2 months on TB treatment, for discriminating between TB cases who converted their sputum to negative by 2 months (early converters) and those who converted after 2 months. (d) Comparison of RISK6 signature scores in IMPRESS participants stratified by detectable (>400 RNA copies/mL plasma) vs. undetectable plasma HIV load (<400 RNA copies/mL plasma). Horizontal lines depict medians, the boxes the IQR and the whiskers the range. Violin plots depict the density of data points. The effect size is the relative difference in median plasma viral load and the p-value was calculated using Mann-Whitney U test.

Although HIV infection causes immunodeficiency, it also drives chronic immune activation and inflammation^18-20^ and induces expression of type I IFN response, including interferon stimulated genes (ISGs)^21,22^. Successful antiretroviral therapy (ART) suppresses viral replication and reduces plasma viral load (pVL), decreasing inflammation and immune activation, although not to levels typical of HIV-uninfected persons^23^. Since RISK6 includes three IFN-inducible ISG transcripts, we aimed to evaluate the effect of pVL on signature scores in the IMPRESS trial. Eighty-five participants had pVL measurements, 36 with detectable viral loads (above 400 copies per mL) and 49 with undetectable viral loads (below 400 copies per mL); sixty of the measurements were baseline samples and 25 were end-of-treatment samples. RISK6 signature scores were significantly higher in samples with detectable pVL than those with undetectable pVL (p = 0.0027, **Figure 5d**), showing that pVL is a confounder in ISG-containing transcriptomic signatures, one that could affect many patients.

### Robustness of the PCR-based RISK6 signature

An advantage of the pair-wise ensemble structure of RISK6 is that a signature score can be calculated even if one or more transcript is not detected, for example due to a failure during PCR amplification. To determine how robust the signature is to such missing data, we compared diagnostic performance for discriminating between HIV-uninfected TB cases and asymptomatic controls (Figure 2) by the full 6-gene RISK6 signature, which comprises nine pairs formed between six transcripts, or after removing one, two, three or four, of these transcripts such that every combination of the pairs was tested. Diagnostic performance was not affected by removal of a single transcript, irrespective of transcript identity (AUC for full RISK6: 93.6, 95%CI 87.4-99.7; average AUC for 5-transcript signature: 93.2, lower 95%CI bound: 85.5%, **Figure 6a**).

**Figure 6.**
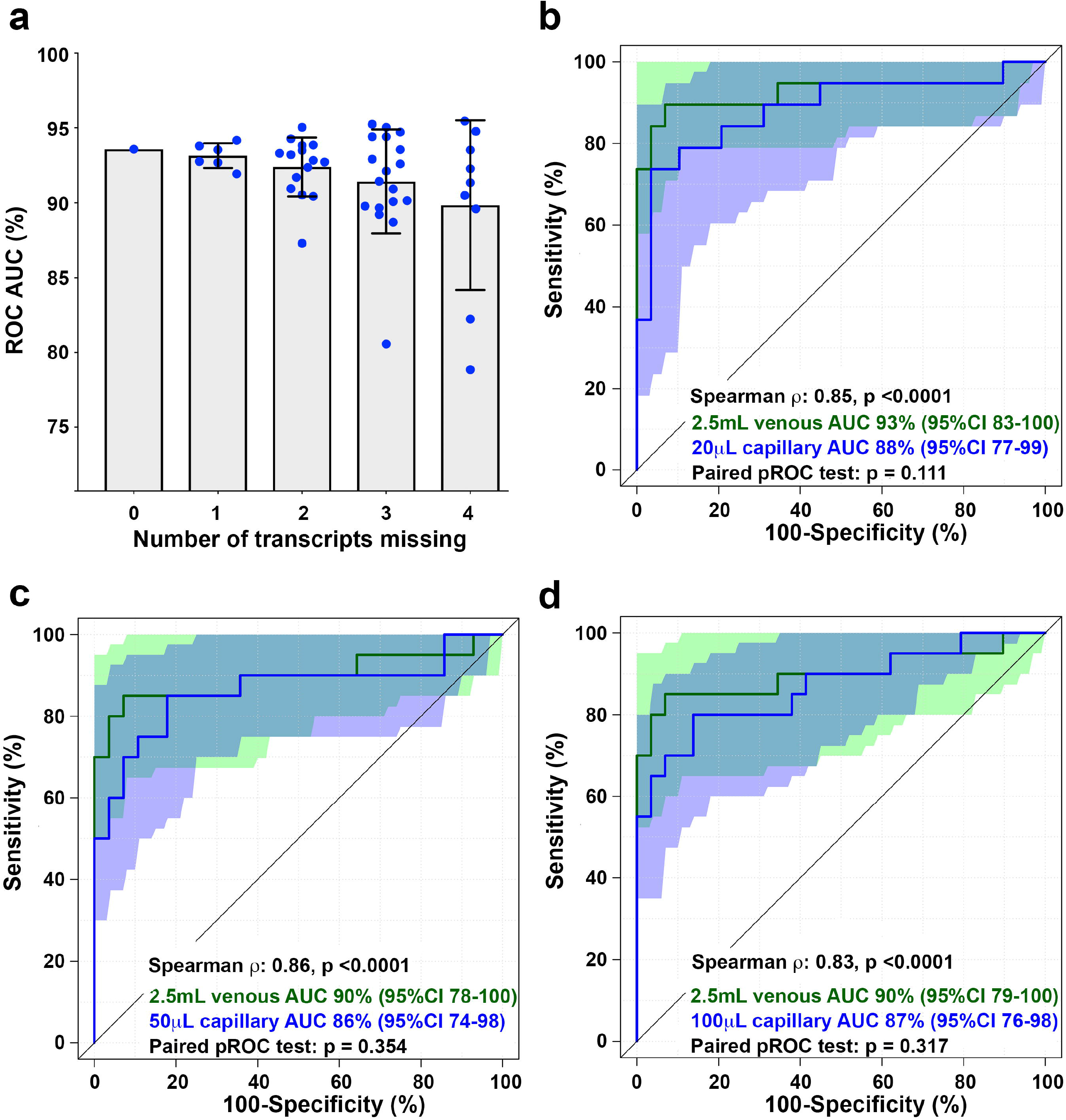
Robustness of RISK6. (a) ROC AUC values for discrimination between HIV-uninfected TB cases and asymptomatic controls in the CTBC cohort by the full 6-gene RISK6 signature (9 pairs formed between 6 transcripts, far left), or after removing 1, 2, 3 or 4 of the transcripts such that every combination of the pairs (represented by individual blue dots) was tested. The grey bar graph represents the mean and the error bar the 95% CI. (b-d) ROC curves depicting performance of RISK6 for discriminating between TB cases and controls in the capillary blood cohort. Green curves represent RISK6 measured on 2.5mL venous blood collected in PAXgene tubes. Blue curves represent RISK6 measured on 20μL (b), 50μL (c) or 100μL (d) capillary blood collected by finger prick. Shaded areas depict the 95% CI. The Spearman correlation coefficients and associated p-values for comparison of RISK6 scores measured by 2.5mL venous blood versus each capillary blood volume are shown. Only individuals with paired venous and capillary blood results were included in each comparison (i.e. venous blood results in those with a missing capillary blood value were excluded from each graph). ROC curves were compared using the pROC function in R and resulting p-values are shown.

However, removal of two or more transcripts, especially when two or more of SERPING1, SDR39U1 or TUBGCP6 were omitted, resulted in somewhat decreased performance of RISK6 (Average AUC for 4-transcript signature: 92.4, lower 95%CI bound: 80.1; average AUC for 3-transcript signature: 91.4, lower 95%CI bound: 72.1). A very similar result was observed when the same analysis was performed on the Brazilian cohort (**Supplementary Figure 3**). These results show that RISK6 can tolerate one or even two missing transcripts without the diagnostic performance being markedly eroded.

Effective deployment of transcriptomic signature tests such as RISK6 in community or primary health care settings is dependent on successful translation of gene expression to methods that are simple, cheap and rapid. An expensive and cumbersome component of any blood transcriptomic assay is the procedure and cost of blood collection. Therefore, we sought to determine if RISK6 could be reliably measured in very small volumes of capillary blood collected by finger stick. We compared discrimination between healthy controls and TB cases by RISK6, measured by qRT-PCR in 20μL, 50μL or 100μL capillary blood, benchmarked against the typical 2.5mL venous blood collected in PAXgene tubes. Among samples collected from the 49 participants, the number of samples with one or more failed PCR reaction, where the amplification curve for one transcript did not pass the QC threshold defined by Fluidigm, for the 20μL, 50μL and 100μL capillary blood volumes was 4 (8%), 3 (6%) and 3 (6%), respectively. None of the 2.5mL venous blood samples yielded failed PCR reactions. When failure of 1 of the 6 transcripts was tolerated, RISK6 scores could be calculated for 98% (1 failed sample), 98% and 100% of the 20μL, 50μL and 100μL capillary blood samples, respectively.

RISK6 signature scores measured on 2.5mL venous blood correlated strongly with those measured on 20μL, 50μL or 100μL capillary blood samples (Spearman rho > 0.83; **Figure 6b-d**). Diagnostic performance of RISK6 was statistically non-inferior when measured in 20μL, 50μL or 100μL capillary blood samples compared with venous blood; ROC analysis yielded equivalent AUC curves (**Figure 6b-d**). These results show that RISK6 can be measured on very small volumes of capillary blood collected by finger stick, which may be amenable to translation to a point of care testing platform.

## DISCUSSION

We discovered and validated RISK6, a parsimonious and robust blood transcriptomic signature with universal applicability for predicting incident TB, as a triage test for identifying individuals with or without respiratory symptoms who should be further investigated for TB disease, and for monitoring the response to TB treatment.

RISK6 identified individuals at risk of progression to incident TB and met or approached the respective benchmarks set out in WHO target product profiles for incipient TB tests^4^. When applied to samples collected within 1 year of TB diagnosis in the ACS discovery cohort, RISK6 met the minimum criteria for a test for progression to TB set out by the WHO and FIND^4^. At a sensitivity of ≥75% a specificity of 82.8% was observed (**Table 2** and **Table 3**). However, when applied to samples collected within 1 year of TB diagnosis in the GC6-74 validation cohort, the specificity at a sensitivity of ≥75% was 50.3%, which did not meet these criteria (**Table 2**). Prognostic performance for incident TB of RISK6 was significantly better than that reported for the previously described 16-gene ACS signature^6^, which was discovered by RNA-seq in the ACS progressor and non-progressor cohort. RISK6 also validated in the independent GC6-74 cohort of TB household contacts by blind prediction. Importantly, performance of RISK6 in distinct cohorts from 3 different African countries was similar, although RISK6 did not significantly discriminate between progressors and non-progressors from Ethiopia, likely due to the small number of progressors, namely 12. The limitations of such small sample sizes for biomarker validation is also evident from other biomarker studies on the GC6-74 cohort. It was notable that the performance of RISK6 in the three GC6-74 cohorts was very similar to the previously published RISK4 signature (**Table 2**), which was specifically developed as a “pan-African signature” ^8^. Recent studies showed that the 16-gene ACS signature, as well as two other small diagnostic signatures^24,25^, did not validate in either one or both of the GC6-74 validation sub-cohorts of Gambian or Ethiopian progressor and non-progressor TB household contacts^8^. However, when the 16-gene ACS signature was measured in the full GC6-74 cohort from The Gambia, comprising 30 progressors and 129 non-progressors^6^, the signature significantly validated by blind prediction. These results highlight the value of longitudinal cohort studies with sufficient incident TB cases to allow reliable assessment of prognostic performance of risk signatures. It is critical that more such cohort studies be performed to increase our collective capacity to develop, refine and validate such biomarkers.

A reliable and simple triage test to identify those who should be investigated more intensively for subclinical or active TB disease is urgently needed to improve case finding strategies and allow earlier diagnosis and treatment. RISK6 also performed well as a triage test in patients with respiratory symptoms who presented for care. However, with 56% specificity at >90% sensitivity, it did not meet the minimum criteria set out in the WHO target product profile (TPP) for a referral test to identify people who may have TB^5^. However, in our post-hoc analyses the specificity of RISK6 in differentiating between definite TB cases and ORD among patients with no prior history of TB was 75% at a set sensitivity of >90% (**Table 3**), which met the WHO target product profile (TPP). Data regarding the interval since the previous TB episode was often unavailable, precluding analysis of this factor. This finding, along with the apparent effect of chest radiographs on RISK6 scores, highlights the importance of including clinical and epidemiological factors in studies of diagnostic biosignatures. Community-based case finding studies and prevalence surveys have shown that a substantial proportion of microbiologically-confirmed TB cases are asymptomatic^26-28^, highlighting the need for TB case finding in asymptomatic communities. RISK6 showed excellent diagnostic performance in differentiating between symptomatic TB cases and asymptomatic controls in four different case-control cohorts from South Africa, Peru and Brazil. Application of RISK6 to the South African cohorts met or exceeded the sensitivity and specificity criteria set out in the TPP for a screening or triage test^5^. In the South American cohorts, however, these criteria were only met when TB cases were compared to uninfected controls as determined by negative QuantiFERON tests. Whether this reflects a real geographic, genetic, environmental or epidemiological difference between South African and South American communities is not clear. For the Peruvian cohort RISK6 measurements were performed on RNA isolated from PBMC, which may have affected diagnostic performance, although we showed that near-identical ROC AUC results were observed when diagnostic performance of the ACS 11-gene signature was measured in whole blood and PBMC^7^. It is noteworthy that diagnostic performance of RISK6 was higher in Brazilian culture+smear+ TB cases (AUC 99.8%, 95%CI 99.4-100) than in culture+smear-TB cases (AUC 90.5%, 95%CI 76.8-100) and that RISK6 scores correlated significantly with lung lesion activity measured by PET in the South African Catalysis cohort. RISK6 scores also decreased during disease resolution upon TB treatment and showed promise as a treatment response biomarker. This reflects the opposite of the increasing inflammatory signals detected by RISK6 during disease progression, as previously reported for transcriptomic signatures^16^. Our findings strongly suggest that disease severity in TB cases play a role in performance of transcriptomic signatures, and likely other biomarkers. Given the lines of evidence that the signature tracks severity of disease and lung lesions, it should be noted that biomarker performance in populations from different settings may be influenced by differences in study design that may preferentially enrol patients with more or less severe disease, rather than reflecting purely geography-associated differences. Larger and well-designed longitudinal biomarker studies are necessary to investigate the performance characteristics of blood biomarkers, such as RISK6, for classifying individuals with ambiguous respiratory phenotypes that are difficult to diagnose, and for revealing which stage of the TB spectrum such individuals may fall into.

Underlying HIV-infection did not significantly affect diagnostic or treatment response performance of RISK6, which is crucial given the high prevalence of undiagnosed TB in people living with HIV^10^. We acknowledge that the effect of HIV was not assessed in all of the validation cohorts and more such analyses are necessary to definitively establish the effects of underlying HIV infection on RISK6 performance. Regardless, other published blood-based transcriptomic TB signatures showed reduced diagnostic performance in HIV-infected compared to uninfected persons^6,26-29^. Since most transcriptomic TB signatures detect the elevation of ISG expression during TB, this effect of HIV is not surprising given that strong Type I IFN responses constitute the typical anti-viral response^29^. Persistent HIV viremia also drives chronic immune activation^30^ which is characterised by high ISG expression. Our data show that HIV-infection was associated with elevated RISK6 signature scores, but that expression levels of individual transcripts in the signature were not dramatically modulated by HIV-infection. Although discrimination between HIV-infected TB cases and controls was not diminished relative to HIV-uninfected people, our results show that a different diagnostic test threshold would be required for HIV-uninfected and HIV-infected populations. A limitation of our analyses of HIV effects is that the clinical studies were not sufficiently powered to investigate the performance of RISK6 in samples with detectable or high pVLs. The issue of limited power will be addressed in a prospective, multicohort study currently underway in South Africa by performing a head-to-head comparison of performance of RISK6 with other signatures, in both HIV-uninfected and HIV-infected persons (clinicaltrials.gov NCT02735590). Our work suggests that underlying HIV infection has a marked effect on performance of IFN response signatures, which requires further examination. Of note, Esmail, Wilkinson and colleagues demonstrated that a transcriptomic TB signature based on complement pathway genes may have greater utility in ART naïve HIV-infected persons^31^. In their study, pVL did not affect circulating immune complexes, which were associated with transcripts involved in the complement pathway.

We found that RISK6 scores correlated significantly with lung lesion activity measured by PET-CT in TB patients of the Catalysis study who underwent TB treatment. RISK6 showed good performance as a treatment response biomarker, decreasing in score during successful treatment and showing very good discrimination between pre-treatment and post-treatment samples in patients with clinical cure, even in patients with underlying HIV-infection. Similar utility as a treatment response biomarker was observed in the Brazilian cohort to that observed in the South African Catalysis cohort. Importantly, in the Catalysis cohort RISK6 significantly predicted treatment failure prior to treatment initiation and differentiated between treatment failures and cured patients with very high accuracy at the end of treatment. These findings suggest that RISK6 detects inflammatory signals associated with the TB disease process in the lungs or other affected sites and that resolution of these processes can be tracked by monitoring gene expression in the blood. Our data provide proof of concept that RISK6 allows treatment monitoring, as has been shown for a number of other transcriptomic signatures^16,32-34^.

We explicitly developed RISK6 with the ultimate objective of translation to a hand-held point-of-care platform and therefore conducted all performance analyses, including the training and all validation cohorts, by qRT-PCR using a standardized protocol and locked-down analysis algorithm. The RISK6 score is computed based on an ensemble of nine transcript pairs using the pair-ratio approach, which uses ratios of transcripts regulated in opposite directions during TB progression, as previously described^8,11,36,37^. This pair-ratio feature of RISK6 eliminates the need for standardisation of gene expression using reference (or housekeeper) transcripts, restricting measurement of the signature to six primer-probes and simplifying data processing steps.

Importantly, RISK6 was measured by a highly standardized, locked-down protocol in our studies. Consequently, RISK6 performance was not subject to the gene expression normalization methods that are typically necessary to overcome reproducibility problems due to sample and batch effects associated with microarray and RNA-sequencing data^35,36^. Regardless, to allow measurement of RISK6 scores in public microarray or RNA-sequencing datasets, we also provide the score computation algorithm, “RISK6geo”, which computes virtually equivalent scores to the RISK6 algorithm from qRT-PCR data. Finally, we showed that RISK6 could be measured on very small volumes of capillary blood collected by fingerstick, with no discernible effect on signature performance.

Our results support work towards incorporation of RISK6 into rapid, capillary-blood-based point-of-care devices for field evaluation in community and primary care settings and implementation studies.

## METHODS

We developed the transcriptomic signature of risk, RISK6, using samples collected from participants of the Adolescent Cohort Study (**Supplementary Figure 1a**). RISK6 was then applied to seven external validation cohorts to determine prognostic and diagnostic performance and utility as a treatment response biomarker (**Supplementary Figure 1b**). Most of these cohorts have been described previously ^6,8,16,37,38^.

### Adolescent Cohort Study (ACS) (RISK6 discovery)

The Adolescent Cohort Study, including selection of progressors and non-progressors, was previously described^6,8,37^. Briefly, among 6,363 healthy adolescents from the Worcester region of the Western Cape, South Africa, who were enrolled, 46 “progressors” were either TST or QuantiFERON TB-Gold In-Tube assay (Qiagen) (QFT)-positive and developed microbiologically-confirmed intrathoracic disease during 2 years of follow-up. Individuals who were TST or QFT-positive at enrolment and remained healthy (no TB disease) during follow-up, and matched the progressors for age, gender, ethnicity, school of attendance and prior history of TB disease, were included as “non-progressors”. Participants were excluded if they developed tuberculosis disease within 6 months of enrolment (or the first TST or IGRA-positive sample) to exclude early asymptomatic disease that could have been present at the time of assessment, or if they were HIV-infected. Longitudinally collected PAXgene samples were available from most participants at six-monthly intervals. The Human Research Ethics Committee of the University of Cape Town approved the study (045/2005) and all participants provided written full informed, while parents or legal guardians provided written, informed consent. All research was performed in accordance with relevant guidelines/regulations.

### GC6-74 Cohort (Prognostic validation)

The Grand Challenges 6–74 project was previously described^6,8,39^. Briefly, HIV-uninfected household contacts of TB cases were longitudinally followed for up to 2 years, with assessments at baseline, at 6 months and at 18 months. TB progressors who developed microbiologically confirmed TB during follow-up were retrospectively identified and matched 1:4 to healthy non-progressors. Individuals in whom TB disease developed within 3 months of baseline were excluded. PAXgene samples collected from 26 Gambian progressors and 116 non-progressors, 41 South African progressors and 164 non-progressors, and 12 Ethiopian progressors and 48 non-progressors were included. Participants provided written, informed consent. Protocols were approved by the Joint Medical Research Council and Gambian Government Ethics Review Committee, Banjul, The Gambia (SCC.1141vs2), the Stellenbosch University Institutional Review Board (N05/11/187) and the Armauer Hansen Research Institute (AHRI) / All Africa Leprosy, TB and Rehabilitation Training Center (ALERT) Ethics Review Committee (P015/10). All research was performed in accordance with relevant guidelines/regulations.

### Cross-sectional TB cohort (CTBC, Diagnostic validation)

Adults with newly diagnosed active TB (sputum Xpert MTB/RIF-positive or liquid culture-positive) were recruited at primary healthcare clinics in Worcester and Masiphumelele, South Africa. Asymptomatic community controls were recruited from the Worcester or Masiphumelele areas. HIV-infection was diagnosed with the Determine HIV1/2 test (Alere). Protocols were reviewed and approved by the Human Research Ethics committee of the Faculty of Health Sciences at the University of Cape Town (HREC 126/2006 and HREC 288/2008). All study participants provided written informed consent and all research was performed in accordance with relevant guidelines/regulations.

In total, 114 HIV-uninfected adults (53 TB cases and 61 asymptomatic controls) and 86 HIV-infected (45 TB cases and 41 asymptomatic controls) were enrolled. Blood was collected in PAXgene tubes at diagnosis in TB cases and at enrolment in asymptomatic controls.

### ScreenTB and AE-TBC cohorts (Diagnostic validation)

Adults aged >18 years who presented at primary health care clinics in Cape Town emergency or medical wards of Tygerberg Hospital in Cape Town with respiratory symptoms compatible with TB, including cough for at least 2 weeks and another symptom including fever, weight loss, hemoptysis or night sweats, were screened for inclusion for the ScreenTB^11^ or the African-European Tuberculosis Consortium (AE-TBC) studies^12,13^. Those who had TB treatment within 90 days, received immunosuppressive medication (ScreenTB) or quinolones or aminoglycosides in the past 60 days (AE-TBC), had a record of alcohol or drug abuse or a haemoglobin level <9g/dL (ScreenTB) or <10g/dL (AE-TBC), or who were pregnant or breastfeeding, where not eligible. HIV-infection was not an exclusion criterion. Using a pre-defined TB classification algorithm^12^ (**Supplementary Table 4**), patients with microbiologically confirmed pulmonary TB were classified as having definite TB. Those with either a single positive sputum smear or with chest radiographs that were compatible with pulmonary TB and who responded to TB treatment were classified as probable TB. Patients whose sputum tested negative, and who were not started on TB treatment were classified as having “other respiratory diseases” (ORD). These ORD patients also did not have a TB diagnosis during 2 months further follow-up. We also performed a post-hoc analysis after unblinding, where a subset of ORD patients with chest radiographs that were compatible with pulmonary TB were analysed as a separate group. All study participants provided written, informed consent. The documents for the ScreenTB and AE-TBC studies were approved by the Health Research Ethics Committee at Stellenbosch University and all research was performed in accordance with relevant guidelines/regulations. Blood was collected in PAXgene tubes at enrolment, before treatment initiation.

### Peruvian Household Contacts Cohort (Diagnostic validation)

Bacillus Calmette-Guérin (BCG)-vaccinated, HIV-uninfected Peruvian participants were recruited through Socios En Salud (SES), an affiliate of Partners in Health from urban and peri-urban settlements around Lima, Peru, as a case-control study. Participants included adults with recently diagnosed microbiologically confirmed, culture-positive, drug-sensitive pulmonary TB disease (active TB, n=48), and clinically asymptomatic household contacts of TB patients assessed within two-weeks of diagnosing the index case. Household contacts were evaluated for signs of TB disease at the time of enrolment, and were excluded if clinical symptoms of TB were present. Healthy household contacts were assessed for M.tb infection using QuantiFERON TB-Gold In-Tube (QFT) assay. Participants with QFT IFNγ responses >= 0.35 international units (IU)/mL were considered latently Mtb infected (QFT-positive, n=49) and uninfected if QFT IFNγ < 0.35 IU/mL (QFT-negative, n=47). Household contacts were evaluated for signs of TB disease at the time of enrolment, and were excluded when clinical symptoms of TB were present. The Institutional Review Board of the Harvard Faculty of Medicine and Partners Healthcare (protocol number IRB16-1173), and the Institutional Committee of Ethics in Research of the Peruvian Institutes of Health approved the study protocol. All adult study participants and parents and/or legal guardians of minors provided informed consent, while minors provided assent. All research was performed in accordance with relevant guidelines/regulations. Peripheral blood mononuclear cells (PBMC) were isolated from 50mL of venous blood using ficoll and cryopreserved at 5X10^6^ cells/cryovial, then shipped to the Brigham and Women’s Hospital for storage. RNA was extracted from 10^6^ cells PBMCs using the RNeasy extraction kit (Qiagen).

### RePORT-Brazil Cohort (Diagnostic validation and treatment response)

Regional Prospective Observational Research for Tuberculosis (RePORT)-Brazil is an ongoing prospective cohort study at five participating centers in Brazil: three in Rio de Janeiro (Instituto Nacional de Infectologia (INI), Clinica de Saude Rinaldo Delmare (Rochina), Secretaria de Saude de Duque de Caxias (Caxias), one in Salvador (Instituto Brasileiro para Investigação da Tuberculose), and one in Manaus (Fundação Medicina Tropical Dr. Heitor Vieira Dourado). RePORT-Brazil enrols participants ≥18 years-old who initiate treatment for culture-confirmed pulmonary TB, and their close contacts. Details of the protocol have been published previously^40-42^. All participants provided written, informed consent and the protocol was approved by the Ethics Committee of the Maternidade Climério de Oliveira, Salvador, Brazil. All research was performed in accordance with relevant guidelines/regulations. Blood was collected in PAXgene tubes at diagnosis in TB cases and at enrolment in contacts.

### Capillary blood cohort

Twenty adults (18 years or older) with recently diagnosed, microbiologically confirmed pulmonary TB, who were positive for either sputum MGIT or solid culture, Xpert MTB/RIF, Xpert MTB/RIF Ultra, or smear microscopy within the preceding two weeks and had received no more than two weeks of tuberculosis treatment were consecutively recruited from ongoing TB diagnostic and treatment studies at the South African Tuberculosis Vaccine Initiative (SATVI) field site. Twenty-nine healthy adults living in communities from the Cape Winelands region were also enrolled. Individuals with anaemia (haemoglobin less than 8.0 g/dl) or any other acute or chronic disease were excluded from both groups but no screening for HIV was performed. For each participant, 2.5mL of venous blood was collected into PAXgene RNA tubes (Qiagen) while 20μl, 50μl or 100μL capillary blood was collected by fingerprick sequentially using 20μl or 50μl Minivettes (Sarstedt) without anti-coagulant and immediately transferred into 0.5mL microtubes (Sarstedt) containing PAXgene fluid at an equivalent ratio to the manufacturer’s recommendations, i.e. 1μl blood: 2.76μl PAXgene fluid. Samples were mixed by inversion (venous PAXgene tubes) or by flicking (capillary blood microtubes), incubated at room temperature for two hours, and stored at −40°C. Participants provided written informed consent and the protocol was approved by the Human Research Ethics committee of the Faculty of Health Sciences at the University of Cape Town (HREC 812/2017). All research was performed in accordance with relevant guidelines/regulations.

### Catalysis treatment response cohort, “Catalysis” (TB treatment response in HIV-uninfected patients)

In total, 131 HIV-uninfected adults with newly diagnosed pulmonary TB, as confirmed by sputum culture, were recruited at primary healthcare clinics in Cape Town; 101 completed the study. Disease pathology was quantified by positron emission tomography and computerized tomography (PET-CT) imaging using ^18^F FDG at baseline, week 4 and week 24. Total glycolytic activity index (TGAI) is a product of lesion volume and FDG uptake intensity and represents the total inflammatory burden, as previously provided^15,16^. PAXgene tubes were collected prior to the start of treatment and at one, four, and 24 weeks after treatment initiation. Of the 101 sample sets sequenced for a transcriptomics analysis^16^, 84 patients met or exceeded the WHO definition for cure after the standard six-month treatment (“cures”, had proven and then maintained sputum culture negativity by month 6). Amongst these, 70 had RNA available for qRT-PCR analysis. Eight patients did not achieve bacteriological cure (classified as “treatment failures”, if the month 6 culture was still positive) and 7 had available RNA. None of the treatment failures achieved culture negativity at any time point during treatment and 7 had RNA for qRT-PCR analysis). The remaining 10 patients were probable cures (only final culture was negative) or unevaluable (treatment response ambiguous) and were not included in any analyses. Twenty-nine healthy controls were also enrolled from the same communities and 21 had RNA available for qRT-PCR analysis. All participants provided written, informed consent and the protocol was approved by the Stellenbosch University Human Research Ethics Committee (N10/01/013). All research was performed in accordance with relevant guidelines/regulations.

### IMPRESS trial cohort (Recurrent TB treatment response in HIV-infected patients)

This study was an open-label, randomized controlled trial, “Improving Retreatment Success” (IMPRESS, clinicaltrials.gov, NTC02114684; SANCTR DOH-27-0414-4576), performed in Durban, KwaZulu-Natal^17^. IMPRESS was designed to determine if a moxifloxacin-containing 24-week regimen, in which moxifloxacin was substituted for ethambutol, would improve TB retreatment outcomes relative to the standard TB treatment regimen. The trial enrolled adults with a previous history of TB disease who received a new diagnosis of drug-sensitive TB by positive Xpert MTB/RIF (Cepheid) or sputum smear microscopy, or both. Sputum samples were collected for culture testing every 2 weeks during the intensive phase of treatment and monthly thereafter until successful treatment completion. Whole blood was collected in PAXgene tubes at baseline, 7 days and 2, 6, 8 and 14 months after start of TB treatment. Sixty-three HIV-infected patients had RNA available and were included in the analyses (44 early converters, with sputum culture conversion before month 2; and 19 late converters, who converted after month 2). The IMPRESS protocol was reviewed and approved by the University of KwaZulu-Natal Biomedical Research Ethics Committee (BREC No. BFC029/13). The IMPRESS trial was also approved by the Medicines Control Council of South Africa (MCC Ref:20130510). All research was performed in accordance with relevant guidelines/regulations.

### RNA extraction

RNA was manually extracted from collected PAXgene Blood RNA tubes (Qiagen) with the PAXgene blood RNA kit (Qiagen) according to the manufacturer’s instructions or on an automated Tecan Freedom EVO 150 robotic platform with the Promega Maxwell SimplyRNA kit, using a modified protocol. Manually extracted RNA was stored at −80°C, and later used for transcriptomic analysis. For RNA extracted by robotic platform an aliquot was immediately used for cDNA synthesis. For the Peruvian cohort, RNA samples were extracted from 10^6^ PBMCs using the RNeasy kit (Qiagen) according to manufacturers’ instructions, and blinded, frozen aliquots of RNA were shipped to the University of Cape Town.

For the venous versus capillary blood comparison, RNA was isolated with the PAXgene blood RNA kit (Qiagen) according to the manufacturer’s instructions with the following modifications: capillary blood samples were washed in 400μl water (instead of 4ml) and homogenised by pipetting to avoid loss of the small pellet; venous and capillary samples were eluted in 80μl and 40μl of PAXgene blood RNA kit elution buffer, respectively.

### Gene expression

cDNA was synthesized from extracted RNA using SuperScript II reverse transcriptase and pre-amplified using a pool of specific TaqMan primer-probe sets for microfluidic qRT-PCR. Gene expression of individual transcripts was then quantified by microfluidic qRT-PCR using either 96.96 or 192.24 Gene Expression chips on the BioMark HD (Fluidigm). An internal positive control sample was run on every chip to monitor inter-chip gene expression consistency.

### Discovery of a parsimonious prognostic signature of TB disease risk

We sought to develop a PCR-based signature comprising a small ensemble of transcript pairs that each represent the ratio between one upregulated and one downregulated transcript in progressors, relative to controls, as described previously^8^. This pair-ratio ensemble format presents two advantages. Firstly, the up-down pairing provides a “self-standardisation” function that eliminates the need for housekeeper transcript-based standardisation of RT-PCR cycle threshold values. Secondly, the ensemble of pairs provides robustness to the signature since a signature score can be calculated even if expression data for one transcript (and its pairs) is not available, due to a failed PCR reaction, for example.

Discovery of RISK6 signature of TB disease risk (**Supplementary Figure 1a**) was performed using all ACS cohort progressor/non-progressor samples collected within 360 days of TB disease diagnosis^6^. We first identified exon junctions that were differentially expressed in RNA-sequencing data from all progressors and matched non-progressors (published in ^6^ and available on GEO: accession number GSE79362). We applied the random subsets approach, which randomly selects a partition of half the samples with a quarter of the features, to train support vector machines of all possible pairs of junctions using the Pair-Ratio approach. The Pair-Ratio approach pairs transcripts that are regulated in opposite directions in progressors and non-progressors. We identified transcript pairs that differentiated progressors and non-progressors with the highest sensitivity and specificity on the remaining partition of samples not used for fitting. This was repeated until pairs that comprised 84 unique exon junctions were identified, such that these could be conveniently assayed, along with 12 housekeeper (reference) transcripts, by microfluidic PCR in a 96-reaction format (**Supplementary Figure 2**).

### Training RISK6, a prognostic PCR signature of TB disease risk

Taqman FAM-TAM primer-probe assays for each of the 84 exon junctions were used to measure expression of all transcripts by microfluidic qRT-PCR using samples from the entire ACS cohort. Delta Ct values were computed for each exon junction relative to the geometric mean of the 12 reference transcripts. To train the best parsimonious signature, we evaluated fit of pair-ratio ensembles consisting of either 10, 8, 6, 4 or 2 transcript pairs and evaluated their ability to differentiate between progressors and non-progressors. An appreciable drop in area under the receiver operating characteristic curve (ROC-AUC) was observed for the 2-transcript and 4-transcript ensembles, compared to the 6-, 8- and 10-transcript pair ratio ensembles (**Supplementary Figure 2)**. As there was no significant difference in performance between the 6-, 8- and 10-transcript models, we selected the 6-primer model, which we termed RISK6, based on performance and smallest functional ensemble size.

### Statistical analysis

The RISK6 signature score is calculated as follows (R-script available on Bitbucket: https://bitbucket.org/satvi/risk6):

1. Measure the cycle thresholds (Cts) for the 6 primer-probe assays listed in **Supplementary Table 1**, by qRT-PCR.
2. For each of the 9 transcript pairs, compute the difference in raw Ct, which produces the log-transformed ratio of expression.
3. Compare the measured ratio to ratios in a look-up table for the given pair of transcripts.
4. Assign a corresponding score in the look-up table to the ratio. If the measured ratio is larger than all ratios in the relevant column of the look-up table, then assign a score of 1 to the ratio.
5. Compute the average over the scores generated from the set of pairs. If any assays failed on the sample, compute the average score over all ratios not including the failed assays. The resulting average is the final score for that sample.

There is considerable interest in the biosignature field to apply such signatures to publicly available microarray or RNA-sequencing data^43^ (Gupta et al., BioRxiv, https://doi.org/10.1101/668137). RISK6 scores can be computed from log_10_-transformed microarray or RNA-sequencing data using the formula:

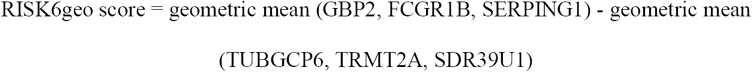

where normalized log_2_-transformed mean fluorescence intensity or normalized read count values of GBP2, FCGR1B, SERPING1, TUBGCP6, TRMT2A and SDR39U1 are used.

RISK6 scores can also be computed using this method from qRT-PCR data using the formula:

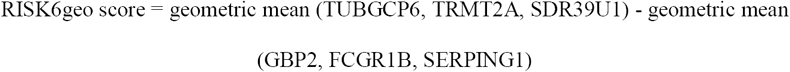

where raw Ct values of GBP2, FCGR1B, SERPING1, TUBGCP6, TRMT2A and SDR39U1 are used. Comparative performance characteristics of the RISK6 and RISK6geo signatures for the different cohorts in this study are shown in **Supplementary Table 2**.

qRT-PCR gene expression data was quality controlled using scripts generated in R and signature scores were calculated. All RISK6 scores, with the exception of those in the discovery cohort, were generated by blinded laboratory personnel. Only once RISK6 score results were locked down and, where appropriate, shared among collaborators, were group allocations unblinded for performance analyses. ROC AUCs were generated and compared using the pROC^44^ and verification^45^ packages in R. Statistical analyses were done using Mann Whitney U for differences between two groups, Wilcoxon ranked sum and Kruskal-Wallis tests for differences between three groups in GraphPad Prism v8. To generate spline plots that show temporal changes in transcript expression between adolescent progressors and non-progressors, we computed log_2_ fold change values between progressor and non-progressors transcript abundance (measured by RNA-sequencing) as previously described^37^, and modeled these as a nonlinear function of TimeToDiagnosis for the entire adolescent progressor/non-progressor cohort using the smooth.spline function in R with three degrees of freedom. Ninety-nine percent confidence intervals for the temporal trends were computed by performing 2000 iterations of spline fitting after bootstrap resampling from the full dataset. The median difference and 95% CIs in expression of RISK6 signature genes was computed from 1000 bootstrapped median Ct values between HIV+ and HIV-individuals. Genes with 95% CI bounds above zero were considered significant.

## Data Availability

The RISK6 scores and associated clinical data for all cohorts are in Supplementary Tables 5-13.

https://bitbucket.org/satvi/risk6

## Data availability statement

The RISK6 scores and associated clinical data for all cohorts are in Supplementary Tables 5-13.

## Author contributions

APN, ET, DZ and TJS conceived the study.

APN, SKM, WAH, SHEK, JW, RW, IVR, BM, MMurray, BBA, TRS, JS, GW, KN, NP, MH, DZ and TJS implemented clinical studies, raised funds and/or provided the resources.

APN, ET, SKM, SCM, SS, NNC, STM, FD, ME, NB, MMurphy, IVR and BBA processed samples, performed the experiments, and analyzed the data.

APN, SS, NNC, NB, MF, MM, CM provided operational and/or project management.

APN, ET, SKM, SCM, SS, NNC, STM, FD, JW, IVR, BM, MMurray, BBA, TRS, GW, MH, DZ and TJS interpreted the results.

APN, ET, SKM, SCM, SS, DZ and TJS wrote the manuscript.

All authors have read and approved the manuscript.

## Consortium and team members

### The Adolescent Cohort Study Team

^1^Fazlin Kafaar, ^1^Leslie Workman, ^1^Humphrey Mulenga, ^1^E. Jane Hughes, ^1^Onke Xasa, ^1^Ashley Veldsman, ^1^Yolundi Cloete, ^1^Deborah Abrahams, ^1^Sizulu Moyo, ^1^Sebastian Gelderbloem, ^1^Michele Tameris, ^1^Hennie Geldenhuys, ^15^Rodney Ehrlich, ^16^Suzanne Verver, ^17^Larry Geiter

### The GC6-74 Consortium

^4^Gillian F. Black, ^4^Gian van der Spuy, ^4^Kim Stanley, ^4^Magdalena Kriel, ^4^Nelita Du Plessis, ^4^Nonhlanhla Nene, ^4^Teri Roberts, ^4^Leanie Kleynhans, ^4^Andrea Gutschmidt, ^4^Bronwyn Smith, ^4^Andre G. Loxton, ^4^Gerhardus Tromp, ^4^David Tabb, ^18^Tom H.M. Ottenhoff, ^18^Michel R. Klein, ^18^Marielle C. Haks, ^18^Kees L.M.C. Franken, ^18^Annemieke Geluk, ^18^Krista E van Meijgaarden, ^18^Simone A Joosten, ^19^W. Henry Boom, ^19^Bonnie Thiel, ^20^Harriet Mayanja-Kizza, ^20^Moses Joloba, ^20^Sarah Zalwango, ^20^Mary Nsereko, ^20^Brenda Okwera, ^20^Hussein Kisingo, ^5^Shreemanta K. Parida, ^5^Robert Golinski, ^5^Jeroen Maertzdorf, ^5^January Weiner 3rd, ^5^Marc Jacobson, ^21^Hazel Dockrell, ^21^Steven Smith, ^21^Patricia Gorak-Stolinska, ^21^Yun-Gyoung Hur, ^21^Maeve Lalor, ^21^Ji- Sook Lee, ^22^Amelia C Crampin, ^22^Neil French, ^22^Bagrey Ngwira, ^22^Anne Ben-Smith, ^22^Kate Watkins, ^22^Lyn Ambrose, ^22^Felanji Simukonda, ^22^Hazzie Mvula, ^22^Femia Chilongo, ^22^Jacky Saul, ^22^Keith Branson, ^1^Hassan Mahomed, ^1^E. Jane Hughes, ^1^Onke Xasa, ^1^Ashley Veldsman, ^1^Katrina Downing, ^1^Humphrey Mulenga, ^1^Brian Abel, ^1^Mark Bowmaker, ^1^Benjamin Kagina, ^1^William Kwong Chung, ^17^Jerry Sadoff, ^17^Donata Sizemore, ^17^S Ramachandran, ^17^Lew Barker, ^17^Michael Brennan, ^17^Frank Weichold, ^17^Stefanie Muller, ^17^Larry Geiter, ^23^Desta Kassa, ^23^Almaz Abebe, ^23^Tsehayenesh Mesele, ^23^Belete Tegbaru, ^24^Debbie van Baarle, ^24^Frank Miedema, ^25^Rawleigh Howe, ^25^Adane Mihret, ^25^Abraham Aseffa, ^25^Yonas Bekele, ^25^Rachel Iwnetu, ^25^Mesfin Tafesse, ^25^Lawrence Yamuah, ^12^Martin Ota, ^12^Philip Hill, ^12^Richard Adegbola, ^12^Tumani Corrah, ^12^Martin Antonio, ^12^Toyin Togun, ^12^Ifedayo Adetifa, ^12^Simon Donkor, ^26^Peter Andersen, ^26^Ida Rosenkrands, ^26^Mark Doherty, ^26^Karin Weldingh, ^27^Gary Schoolnik, ^27^Gregory Dolganov, ^27^Tran Van

### The ScreenTB Consortium

^4^Petri Ahlers, ^4^Gian van der Spuy, ^4^Ilana van Rensburg, ^4^Hygon Mutavhatsindi, ^4^Portia Manngo, ^4^Kim Stanley, ^4^Andriette Hiemstra, ^4^Shirley McAnda, ^12^Joseph Mendy, ^12^Awa Gindeh, ^12^Georgetta Mbayo, ^12^Ebrima Trawally, ^12^Olumuyiwa Owolabi, ^20^Harriet Mayanja-Kizza, ^20^Mary Nsereko, ^20^Anna-Rita Namuganga, ^20^Saudah Nambiru Kizito, ^25^Adane Mihret, ^25^Sosina Ayalew, ^25^Rawleigh Howe, ^25^Azab Tarekegne, ^25^Bamlak Tessema, ^28^Emmanuel Nepolo, ^28^Joseph Sheehama, ^28^Gunar Gunther, ^28^Azaria Diergaardt, ^28^Uapa Pazvakavambwa, ^21^Hazel Dockrell, ^18^Tom Ottenhoff, ^18^Elisa Tjon Kon Fat, ^18^Shannon Herdigein, ^18^Paul Corstjens, ^18^Annemieke Geluk

### The AE-TBC Consortium

^4^Magdalena Kriel, ^4^Gian van der Spuy, ^4^Andre G. Loxton, ^4^Kim Stanley, ^4^Belinda Kriel, ^4^Leigh A Kotzé, ^4^Dolapo O. Awoniyi, ^4^Elizna Maasdorp, ^12^Olumuyiwa Owolabi, ^12^Abdou Sillah, ^12^Joseph Mendy, ^12^Awa Gindeh, ^12^Simon Donkor, ^12^Toyin Togun, ^12^Martin Ota, ^20^Harriet Mayanja-Kizza, ^20^Ann Ritah Namuganga, ^20^Grace Muzanye, ^20^Mary Nsereko, ^20^Pierre Peters, ^28^Marieta van der Vyver, ^28^Faustina N Amutenya, ^28^Josefina N Nelongo, ^28^Lidia Monye, ^28^Jacob A Sheehama, ^28^Scholastica Iipinge, ^22^Amelia C Crampin, ^22^Felanji Simukonda, ^22^Alemayehu Amberbir, ^22^Femia Chilongo, ^22^Rein Houben, ^23^Desta Kassa, ^23^Atsbeha Gebrezgeabher, ^23^Getnet Mesfin, ^23^Yohannes Belay, ^23^Gebremedhin Gebremichael, ^23^Yodit Alemayehu, ^25^Rawleigh Howe, ^25^Adane Mihret, ^25^Yonas Bekele, ^25^Bamlak Tessema, ^25^Lawrence Yamuah, ^18^Tom H.M. Ottenhoff, ^18^Annemieke Geluk, ^18^Kees L.M.C. Franken, ^18^Paul L.A.M. Corstjens, ^18^Elisa M. Tjon Kon Fat, ^18^Claudia J. de Dood, ^18^Jolien J. van der Ploeg-van Schip, ^26^Ida Rosenkrands, ^26^Claus Aagaard, ^5^Maria M. Esterhuyse, ^21^Jacqueline M. Cliff, ^21^Hazel M. Dockrell

### The RePORT Brazil Team

^10^Bruno B. Andrade, ^10^Juan M. Cubillos-Angulo, ^10^Kiyoshi F. Fukutani, ^10^Laise Paixão, ^10^Ricardo Khouri, ^10^Sayonara Melo, ^10^Alice Andrade, ^10^Jéssica Rebouças-Silva, ^10^Hayna Malta, ^10^Artur T. L. Queiroz, ^29^Valeria C. Rolla, ^29^Solange Cavalcante, ^30^Betina Durovni, ^31^Marcelo Cordeiro-Santos, ^32^Afranio Kritski, ^32^José R. Lapa e Silva, ^11^Marina C. Figueiredo

### Peruvian Household Contacts Cohort Team

^3^Kattya Lopez Tamara, ^33^Kattya Lopez Tamara, ^33^Segundo R León, ^33^Leonid Lecca Garcia

### The CAPRISA IMPRESS team

^13,14^Dhineshree Govender, ^13,14^Razia Hassan-Moosa, ^13,14^Anushka Naidoo, ^13,14^Rochelle Adams, ^13,14^Natasha Samsunder, ^13,14^Lara Lewis

### Affiliations

^15^School of Public Health and Family Medicine, University of Cape Town, Cape Town, South Africa

^16^KNCV Tuberculosis Foundation, The Hague, and Amsterdam Institute of Global Health and Development, Academic Medical Centre, Amsterdam, The Netherlands

^17^Aeras, Rockville, MD, USA

^18^Department of Infectious Diseases, Leiden University Medical Centre, Leiden, The Netherlands

^19^Tuberculosis Research Unit, Department of Medicine, Case Western Reserve University School of Medicine and University Hospitals Case Medical Center, Cleveland, Ohio, USA

^20^Department of Medicine and Department of Microbiology, College of Health Sciences, Faculty of Medicine, Makerere University, Kampala, Uganda

^21^Department of Immunology and Infection, Faculty of Infectious and Tropical Diseases, London School of Hygiene & Tropical Medicine, London, United Kingdom

^22^Karonga Prevention Study, Chilumba, Malawi

^23^Ethiopian Health & Nutrition Research Institute, Addis Ababa, Ethiopia

^24^University Medical Centre, Utrecht, The Netherlands

^25^Armauer Hansen Research Institute, Addis Ababa, Ethiopia

^26^Department of Infectious Disease Immunology, Statens Serum Institute, Copenhagen, Denmark

^27^Department of Microbiology and Immunology, Stanford University, Stanford, California, USA

^28^University of Namibia, Windhoek, Namibia

^29^Instituto Nacional de Infectologia Evandro Chagas, Fundação Oswaldo Cruz, Rio de Janeiro, Brazil

^30^Secretaria Municipal de Saúde do Rio de Janeiro, Coordenação de Doenças Transmissíveis, Rio de Janeiro, Brazil

^31^Fundação de Medicina Tropical Doutor Heitor Vieira Dourado, Manaus, Brazil

^32^Hospital Universitário Clementino Fraga Filho, Universidade Federal do Rio de Janeiro, Rio de Janeiro, Brazil

^33^Socios En Salud, Lima, Peru

